# Remote ambulation monitoring enhances composite disability measurement in MS to deliver reduced trial sample size

**DOI:** 10.64898/2026.09.03.26361524

**Authors:** Andrea Festanti, Dimitar Stanev, Cedric Simillion, João Rodrigues, Letizia Leocani, Agne Kazlauskaite, James Overell, Johannes Lorscheider, Gary Cutter, Helmut Butzkueven, Licinio Craveiro, Mike D Rinderknecht

**Affiliations:** F. Hoffmann-La Roche Ltd., Basel, Switzerland; Faculty of Medicine, University Vita-Salute San Raffaele, Milan, Italy; Scientific Institute IRCCS San Raffaele, Milan, Italy; Reha Rheinfelden, Rheinfelden, Switzerland; Department of Neurology, University Hospital Basel and University of Basel, Basel, Switzerland; Department of Biostatistics, University of Alabama at Birmingham, Birmingham, AL, USA; Department of Neuroscience, School for Translational Medicine, Monash University, Melbourne, Australia; Department of Neurology, Alfred Health, Melbourne, Australia

## Abstract

In multiple sclerosis trials, event-based endpoints like the composite confirmed disability progression (cCDP) are often limited by infrequent in-clinic testing. We present a framework that converts high-frequency, remote smartphone-based gait assessments into progression events, and establishes a new composite endpoint that blends digital and clinical progression events. Evaluated in the Phase 3b CONSONANCE study (N = 667), this digitally enhanced endpoint reduced required trial sample size by 24.9% at Week 48 compared to the original cCDP. Furthermore, it achieved equivalent statistical power 24 weeks earlier (Week 72 vs Week 96), demonstrating potential to shorten trial observation periods by 25%. In conclusion, this framework provides a practical pathway to optimize clinical trial efficiency in MS and other neurological conditions, potentially reshaping the path to therapeutic innovation.

## Introduction

Multiple sclerosis (MS) clinical trials increasingly utilize composite endpoints such as the composite confirmed disability progression (cCDP) which is defined by occurrence of a first event of progression in either Expanded Disability Status Scale (EDSS), 9-Hole Peg Test (9HPT), or Timed 25-Foot Walk (T25FW), that has to be confirmed at a subsequent visit. ^1^ ^2^ ^3^ By incorporating different measurement modalities, cCDP improves sensitivity to detect disability progression compared to using EDSS alone. ^1^ However, these are infrequent in-clinic assessments which limits their ability to capture early disease progression or the full spectrum of disease dynamics in a real-world environment. Furthermore, episodic clinic visits are susceptible to measurement variability and increase the risk of regression to the mean, which can obscure true changes or result in false positive progression events.

Digital health technologies, such as wearable sensors or smartphones, can help address these limitations by enabling frequent, highly reliable gait assessments outside the clinical setting. These tools provide a detailed evaluation of continuous real-world ambulatory performance, capturing metrics related to gait quality (i.e., how people walk), gait quantity (e.g., speed or distance people can walk), and perceived walking safety during walking. ^4^ Higher frequency measurements can allow an averaging of random, unpredictable errors thus improving signal-to-noise ratio. ^5^ Integrating these digital metrics into composite endpoints therefore offers a clear pathway to optimize their sensitivity and specificity, enable earlier detection of progression events, and ultimately improve clinical trial efficiency.

This study introduces a novel framework to monitor gait impairment in patients with MS which uses the concept of gait worsening events using remote, unsupervised digital measures. We also assess if a new composite endpoint that blends clinical and digital progression events can outperform the traditional clinical endpoint by reducing sample size requirements and/or achieve the required statistical power earlier in a study (**Figure 1**).

**Figure 1.**
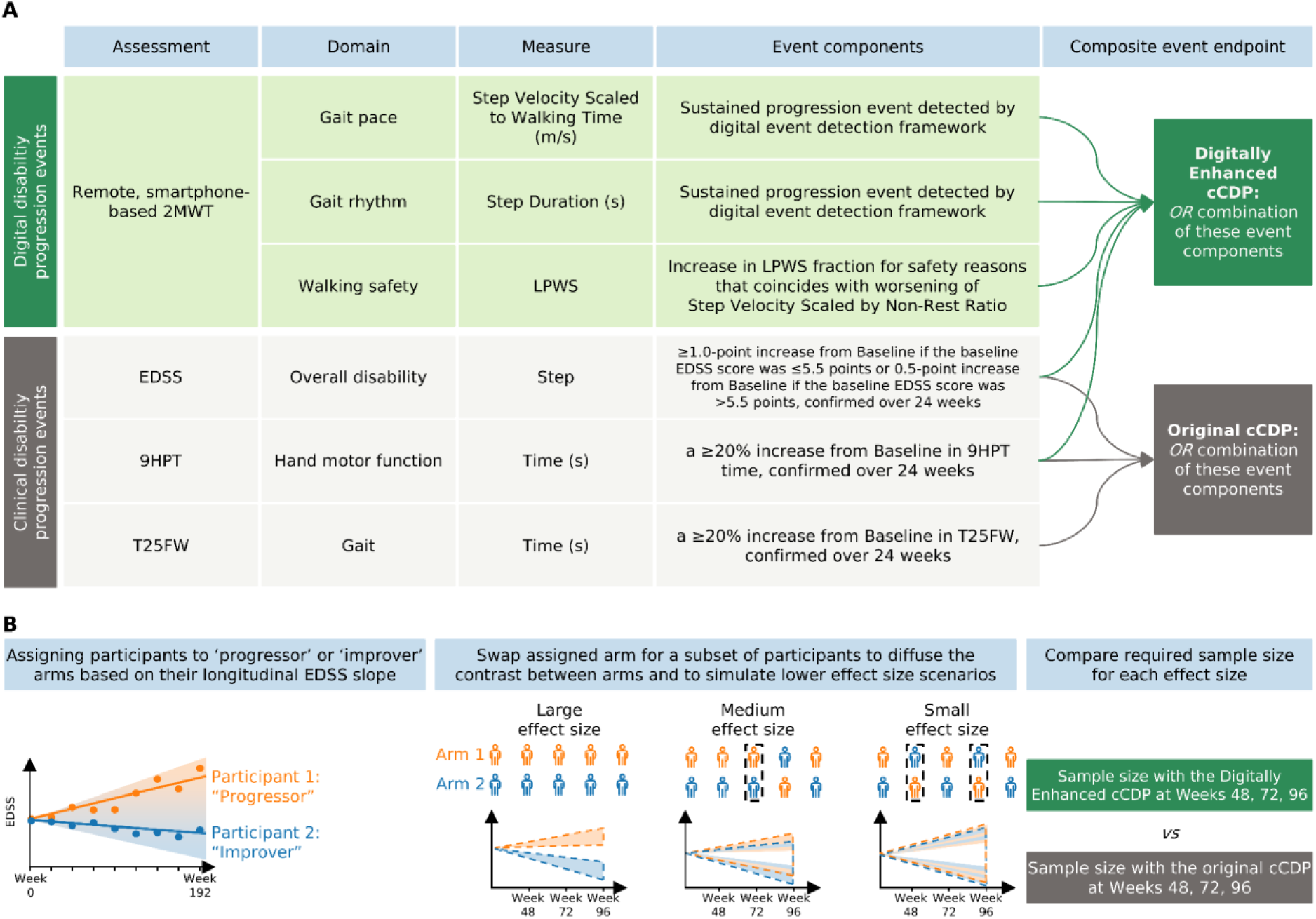
Framework for evaluating progression-based endpoints. (A) Composite confirmed disability progression (cCDP) was defined as a progression event in any of the following: EDSS, 9HPT, or T25FW (logical OR). For the Digitally Enhanced cCDP, the T25FW component was replaced by progression events on digital gait measures derived from remote, smartphone-based 2MWT. These digital measures include Step Velocity Scaled to Walking Time (i.e., step velocity scaled by the proportion of walking bouts during a 2MWT), Step Duration, and LPWS (i.e., the fraction of 2MWT that were skipped by the participant because they felt it was unsafe for them to complete the test). (B) To evaluate the ability of the Digitally Enhanced cCDP to reduce sample size in future clinical trials, the single-arm CONSONANCE cohort was stratified into “progressors” (EDSS increase of ≥0.5 points over 4 years) and “improvers” (EDSS decrease of ≥0.5 points over 4 years). To simulate different effect sizes that could be encountered in clinical trials, small, medium, and large effect sizes were modeled by stochastically reassigning participants between arms. Finally, the required sample sizes to detect these effect sizes with a statistical power of 80% were calculated for both the Digital Enhanced and original cCDP and for different simulated study durations/ visits (48, 72, and 96 weeks). 2MWT, Two-Minute Walk Test; 9HPT, Nine-Hole Peg Test; cCDP, composite Cumulative Disability Progression; EDSS, Expanded Disability Status Scale; LPWS, Loss of Perceived Walking Safety; T25FW, Timed 25-Foot Walk.

While this framework demonstrates the value of validated digital gait measures, it also establishes a foundation for incorporating multi-domain digital metrics, which can help refine the design of future clinical trials in MS and other neurological conditions.

## Methods

In this study we propose a framework to monitor gait function using a remote, unsupervised smartphone-based gait test (i.e. 2-minute walk test) which mimics the concept of disability progression events typically derived from clinical measures (e.g. timed 25-foot walk test) in MS clinical trials. We then assess whether a new composite endpoint that blends clinical and digital progression events can outperform the traditional clinical endpoint by reducing sample size requirements and/or achieve the required statistical power earlier in a study.

### Data Sources

This study utilized data from two distinct cohorts: GaitLab study (ISRCTN15993728) ^6^ and the CONSONANCE trial (NCT03523858) ^7^. The GaitLab study was an observational cohort of individuals with MS (n = 100) and healthy controls (n = 35) who performed, over a 2-week period, daily remote gait assessments (2-Minute Walk Test) using a provisioned smartphone-solution. Participants were instructed to walk as fast as possible, but safely, while avoiding sharp turns for 2 minutes. They were allowed to use their usual walking aid and/or orthotic or rest if needed during the 2MWT. Data from this study was previously used to validate the gait processing pipeline, ^8^ and to develop and select the digital gait metrics with the most robust psychometric properties. In the present work, data were used to quantify measurement noise and determine the Minimal Detectable Change (MDC) of the selected digital gait metrics.

CONSONANCE is an ongoing Phase 3b, multicenter, open-label, single-arm study evaluating the effectiveness and safety of ocrelizumab in adults with primary or secondary progressive multiple sclerosis (MS) and an EDSS score ≤6.5. Clinical functional assessments, including the EDSS, T25FW test, and 9HPT, were administered in-clinic every 24 weeks up to Week 192. In addition, participants performed the same daily, unsupervised, remote walking test as in GaitLab (2MWT) while carrying a provisioned smartphone around the waist in a belt bag. Data from CONSONANCE were used to assess the performance of the new composite endpoint combining digital and clinical progression events. The studies were approved by the relevant local institutional review boards or ethics committees. All patients provided written informed consent on dedicated forms (ICFs).

### Conceptual Development of Digital Gait Progression Events

Clinical progression events in a trial are usually defined as a worsening in clinical status or performance between baseline and a given clinical visit. The degree of worsening must fulfil a minimum threshold which is considered clinically meaningful, and it must be confirmed at a subsequent visit to reduce the risk of capturing random disease fluctuations. For example, a confirmed progression event on T25FW is usually defined as a ≥20% increase from baseline in the time to walk 25-feet as quickly as possible, which is still sustained typically 12 or 24 weeks later. ^9^ A similar framework is proposed here to develop digital gait progression events.

#### Digital Gait Measures

To comprehensively evaluate ambulatory performance during the remote 2MWT, three complementary digital measures were derived: two continuous measures of gait mechanics and one categorical measure reflecting safety perceptions. Step Velocity Scaled to Walking Time (m/s) and Step Duration (s) were extracted from smartphone sensor data using a state-of-the-art adaptive step-detection algorithm. ^8^ Step Velocity Scaled to Walking Time (hereafter referred to as Step Velocity) quantifies overall gait pace adjusted for non-walking rest periods (patients were allowed to rest during the 2MWT, if needed) where lower values indicate worse performance. Step Duration measures gait rhythm, where larger values correspond to longer step execution times and greater impairment. Both measures are highly relevant to the patient and show excellent psychometric properties (**Supplementary Table S1**). To complement these continuous parameters, Loss of Perceived Walking Safety (LPWS) was defined as a categorical metadata measure. Prior to initiating each 2MWT execution, participants answered a mandatory safety prompt (“*Can you walk safely for 2 minutes?*”). LPWS was calculated as the proportion of tests skipped specifically due to a negative safety response within a given window, capturing behavioral indicators of worsening mobility that might precede measurable declines on speed-based measures.

#### Confirmation and Aggregation Windows

To convert high-frequency, remote digital data into reliable progression endpoints, daily measurements were aggregated over structured time intervals to differentiate genuine clinical deterioration from transient performance fluctuations. In CONSONANCE, clinical assessments were performed once every 24 weeks whereas for the smartphone-based 2MWT patients were instructed to execute the test daily. As a result, for the digital gait measures, additional steps for data aggregation are taken. First, a minimum number of completed digital tests (n = 3) was required within an aggregation window to calculate the aggregated measure. This rule ensures that a single anomalous test or outlier does not unduly influence the results, and previous analyses have shown that aggregating across 2 or more tests is sufficient to achieve excellent test-retest reliability. ^10^ The length of these aggregation windows was tailored to each digital measure according to their psychometric properties: Step Velocity utilized 4-week aggregation windows, whereas Step Duration utilized shorter 2-week aggregation windows, owing to its superior test-retest reliability (**Supplementary Table S1**). To confirm that an observed change reflected sustained disease progression, the deterioration had to persist across a predefined 12-week confirmation period. This confirmation period therefore comprised three consecutive 4-week sub-windows for Step Velocity and six consecutive 2-week sub-windows for Step Duration. Sensitivity analyses employed a 24-week confirmation window (in line with clinical confirmation windows), meaning that six consecutive 4-week sub-windows were used for Step Velocity, and twelve 2-week sub-windows were used for Step Duration.

#### Thresholds for Change

To ensure that identified digital progression events represented both statistically robust and clinically meaningful changes, candidate progression events were evaluated against dual criteria: the Minimal Detectable Change (MDC) and the Minimal Clinically Important Difference (MCID).

The MDC threshold was established to account for measurement variability and noise relative to patient adherence (as a function of *n* completed tests within a window). The MDCs were calculated according to the equation (1)

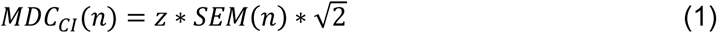

where *z* is the *z*-score for the desired confidence level (e.g., 1.96 for 95%, 2.24 for 97.5%, or 2.58 for 99%), and *SEM* is the Standard Error of Measurement. ^11^ ^12^ To account for the varying number of 2MWTs per participant in the GaitLab dataset, a random-intercept repeated-measures model was used to estimate the *SEM* for *n* median-aggregated tests. The MDC was computed for up to seven aggregated tests (*n* ≤ 7) and parametrized as a decaying power function.

For Step Velocity, the variable MDC threshold was defined as *_MDC_*_97.5_*(n)* = 0.4288 ⋅*n*^−0.4936^ m/s, truncating at a fixed floor of 0.1641 m/s for *n* ≥ 7. For Step Duration, a stricter variance threshold was applied: *_MDC_*_99_*(n)* = 0.1964 ⋅ *n*^−0.5989^ s, capping at a fixed floor of 0.0613 s for *_n_* ≥ 7 (**Supplementary Figure S1**). Concurrently, previously determined anchor-based MCID progression thresholds ^13^ (decrease of ≥ 0.18 m/s for Step Velocity Scaled to Walking Time and an increase of ≥ 0.06 s for Step Duration) were applied to guarantee clinical relevance. True digital progression required an absolute worsening to surpass both the adherence-adjusted MDCs and the MCID threshold across all sub-windows within the 12-week confirmation period.

To operationalize a progression event for the measure Loss of Perceived Walking Safety, we developed a heuristic threshold based on baseline clinical observations. A progression event was defined by two concurrent criteria within an aggregation window: an increase in skipped tests due to safety concerns exceeding 20% (up from a baseline of less than 20%), combined with a simultaneous decrease in continuous Step Velocity.

**Figure 2** illustrates a practical example of a patient with a confirmed progression event on Step Velocity and **Table 1** summarizes the parameters used in the framework to detect gait progression events.

**Figure 2.**
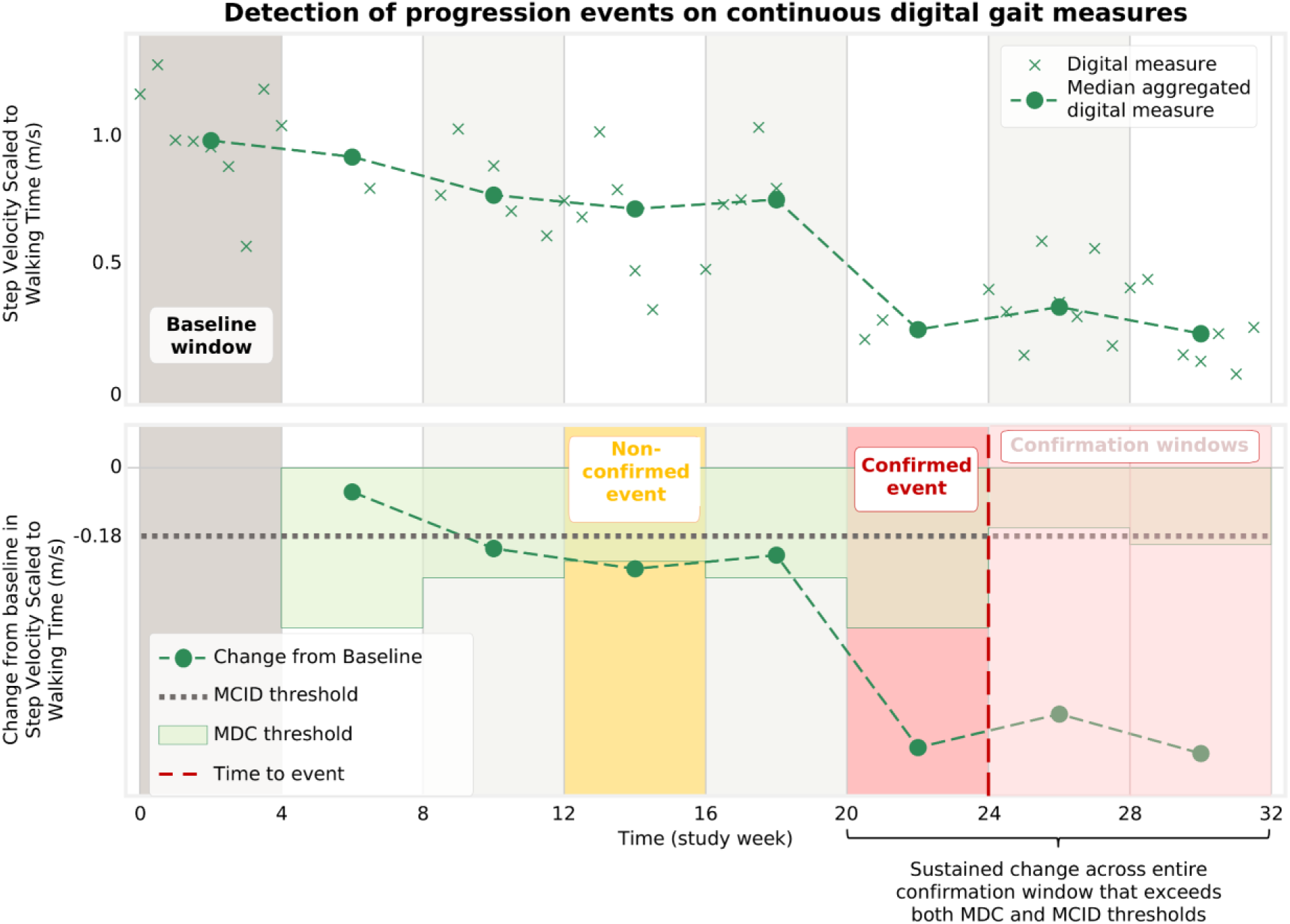
Illustrative example of defining progression events on continuous digital measures. The top plot shows the obtained digital measures from multiple tests and median aggregations in consecutive 4-week windows, whereas the bottom plot shows the change from baseline of these aggregated values. An aggregation window size (e.g., 28 days) is chosen based on clinical insights and patient adherence, with digital measures within each window typically aggregated using the median. The horizontal purple dotted line and the green bars indicate the two thresholds: MCID for the respective measure, and the MDC, which varies depending on the minimum amount of data points available in either the baseline window or the respective window (given that both are required to compute a change). The change from baseline corresponding to the week 12-16 shows an example where no event is detected, due to the subsequent change magnitude not exceeding the MDC threshold. In contrast, the change in the window 20-24 could be confirmed in the subsequent windows (assuming 2 confirmation windows), hence triggering a progression event aligned to the end of the first window (i.e., red dashed line at week 24).

**Table 1.** Parameters used in the framework to detect gait progression events on continuous digital gait measures.

|  | Step Velocity Scaled to<br>Walking Time | Step<br>Duration |
| --- | --- | --- |
| Confirmation window length | 12 weeks | 12 weeks |
| Aggregation window length | 4 weeks | 2 weeks |
| Number of aggregation windows<br>within a confirmation window | 3 | 6 |
| Minimum number of tests per<br>window | 3 | 3 |
| Change thresholds (unit) | (m/s) | (seconds) |
| MDC for < 7 tests within an<br>aggregation window<sup>a</sup> | $MDC_{97.5}(n) = 0.4288 * n^{-0.4936}$ | $MDC_{99}(n) = 0.1964 * n^{-0.5989}$ |
| MDC for ≥7 tests within an<br>aggregation window<sup>a</sup> | $MDC_{97.5} = 0.1641$ | $MDC_{99} = 0.0613$ |
| MCID | -0.18 | +0.06 |

<sup>a</sup>MDCs were derived from the GaitLab study; <sup>6</sup> baseline demographics and disease characteristics of this study are reported in **Supplementary Table S2**; MDC, minimal detectable change; MCID, minimal clinically important difference.

### Comparing Performance Between Conventional and Digitally Enhanced Composite Endpoints

#### Composite Confirmed Disability Progression

To establish a benchmark for comparison, the conventional composite Confirmed Disability Progression (cCDP) was defined according to standard clinical trial criteria. An initial clinical progression event was defined by the occurrence of any of the following: either an EDSS increase of ≥ 1.0 points if the baseline EDSS score was ≤5.5 points or ≥0.5 points if the baseline EDSS score was > 5.5 points, or an increase of ≥20% in T25FW or 9HPT scores. A clinical event was considered confirmed (cCDP) if the worsening was sustained at a subsequent clinical visit at least 24 weeks later. The overall time-to-event for the original cCDP was defined as the earliest confirmed event across any of the three clinical components (EDSS, T25FW, or 9HPT).

For the Digitally Enhanced cCDP, the conventional T25FW component was replaced by digital gait progression events derived from the smartphone-based 2MWT (Step Velocity, Step Duration, or LPWS), while retaining the clinical EDSS and 9HPT components. Time-to-event for the Digitally Enhanced cCDP was defined as the earliest confirmed event among EDSS, 9HPT, or any of the digital gait progression criteria.

#### Simulation of a Two-arm Clinical Trial

Because CONSONANCE is a single-arm study, a two-arm clinical trial was simulated by stratifying participants into “progressor” and “improver” cohorts based on their long-term EDSS trajectories over 4 years (192 weeks). Given that disability progression in this population is heavily driven by ambulatory decline, long-term EDSS slope served as a proxy for true underlying progression speed.

A linear regression model was fitted to all longitudinal EDSS scores from baseline through Week 192 for each participant. Participants with an EDSS change slope of ≥ +0.5 points over 4 years (representing the minimal clinically detectable step on the EDSS scale) were assigned to the progressor arm. Conversely, participants with an EDSS slope of ≤ -0.5 points over the same period were assigned to the improver arm. This artificial contrast created a clear ground-truth benchmark to evaluate how effectively each endpoint distinguished true progressors from improvers. Participants who dropped out or were censored prior to a given landmark visit (Weeks 48, 72, or 96) were excluded from that specific duration analysis.

#### Survival Analyses and Sample Size Modeling

To compare endpoint sensitivity and timing, Kaplan-Meier survival curves were estimated for both the progressor and improver arms, defining event-free survival as the absence of a confirmed progression event. Separation between the survival curves of both arms was assessed to evaluate both the magnitude and onset of differentiation achieved by individual digital measures versus traditional clinical components.

To evaluate required sample sizes for a balanced, two-arm clinical trial, the larger cohort arm was down-sampled without replacement to match the size of the smaller arm. This balanced sampling procedure was repeated across 200 bootstrap iterations. For each iteration, Cox Proportional Hazards models ^14^ were fitted to estimate the hazard ratio (HR) between progressor and improver arms for both the original and Digitally Enhanced cCDP endpoints.

Using Freedman’s method, ^15^ the sample size required to detect these hazard ratios with 80% statistical power (1 − *_β_* = 0.80) and a two-sided significance level of *_α_* = 0.05 was calculated for both endpoints at Weeks 48, 72, and 96. Relative sample size reduction was expressed as the percentage change in required participants using the Digitally Enhanced cCDP compared with the original cCDP.

To test whether performance gains generalized to typical clinical trial scenarios with smaller treatment effect sizes, sensitivity analyses were performed. Effect sizes were systematically attenuated by stochastically swapping a fixed number of participants (*_k_* = 0,10,20,30) between the progressor and improver arms within each bootstrap iteration. Sample size calculations and hazard ratios were recalculated across all values of *_k_* to confirm that the efficiency gains of the Digitally Enhanced cCDP remained robust across varying treatment effect magnitudes.

## Results

### Study Cohort and Arm Stratification

The CONSONANCE cohort evaluated in this analysis comprised 667 individuals with progressive MS with digital data collection starting at baseline (**Table 2**). Baseline characteristics reflected a representative progressive MS population (median EDSS score: 5.5; 56.8% secondary progressive, 43.2% primary progressive). Based on 4-year longitudinal EDSS trajectory slopes, participants were stratified into “progressor” (≥ +0.5 EDSS points over 4 years; n = 237, 236, and 234 at Weeks 48, 72, and 96) and “improver” (≤ -0.5 EDSS points; n = 86, 84, and 78 at corresponding weeks) arms to evaluate endpoint performance in a simulated two-arm trial.

**Table 2.** Baseline demographics and disease characteristics of patients in the CONSONANCE trial

|  | **N = 667<sup>a</sup>** |
| --- | --- |
| **Age**, years, median (IQR) | 51.0 (43.0 – 57.0) |
| **Female**, n (%) | 360 (54) |
| **Type of MS**, n (%) |  |
| Secondary progressive MS | 379 (56.8) |
| Primary progressive MS | 288 (43.2) |
| **Time since MS symptom onset**, years, median (IQR) | 10.6 (6.1 – 16.4) |
| **T25FW**, s, median (IQR) | 9.0 (6.2 – 15.5) |
| **Ambulation Score (EDSS)**, median (IQR) | 4.0 (1.0 – 6.0) |
| **MSWS-12 v2 Transformed Total Score**, median (IQR) | 68.8 (43.8 – 83.3) |
| **EDSS** |  |
| Median (IQR) | 5.5 (4.0 – 6.0) |
| 0 to 3.5, n (%) | 144 (22) |
| 4.0 to 5.5, n (%) | 222 (33) |
| 6.0 to 6.5, n (%) | 301 (45) |

<sup>a</sup>Including all PwPMS with digital gait data collection starting at baseline; 10 PwPMS were censored between Weeks 48 and 72, and 12 PwPMS between Weeks 48 and 96. EDSS, Expanded Disability Status Scale; MS, multiple sclerosis; MSWS-12, 12-item Multiple Sclerosis Walking Scale; PwPMS, people with progressive multiple sclerosis; T25FW, Timed 25-Foot Walk.

### Digital Progression Events

Applying the digital event framework demonstrated distinct separation between progressor and improver arms across all three digital components: Step Velocity, Step Duration, and Loss of Perceived Walking Safety (all Cox *_p_* < 0.0001). Crucially, events were virtually absent in the improver arm (including zero events for LPWS), indicating that the framework effectively filters out measurement noise and eliminates false-positive progression signals in individuals showing EDSS improvement (**Figure 3**).

**Figure 3.**
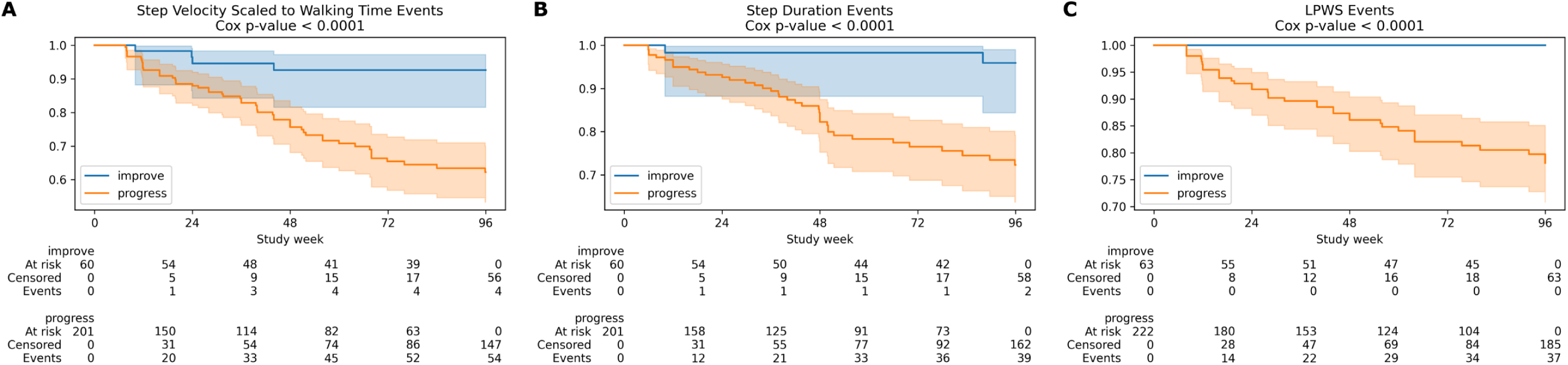
Survival curves showing progression events for each digital gait measure as split by EDSS improvers and progressors. Survival curves are shown for Step Velocity Scaled to Walking Time (A), Step Duration (B), Loss of Perceived Walking Safety (C). Participants with an EDSS change slope of ≥ +0.5 points over 4 years (representing the minimal clinically detectable step on the EDSS scale) were assigned to the progressor arm. Conversely, participants with an EDSS slope of ≤ -0.5 points over the same period were assigned to the improver arm. The sizes of the improver and progressor arms vary due to differences in test adherence and data availability at the different time points.

### Rate and Timing of Progression Events in Digitally Enhanced cCDP versus original cCDP

Substituting the in-clinic T25FW with digital gait events resulted in a similar total proportion of progression events in the progressor arm, with 61.1% for Digitally Enhanced cCDP vs. 62.4% for original cCDP at Week 96 (**Table 3**). However, the Digitally Enhanced cCDP showed less false-positive events in the improver arm (14.1% vs. 16.7%). Furthermore, among participants who experienced events detected by both endpoints (n = 115), the Digitally Enhanced cCDP identified progression earlier in 42.6% of cases, compared to 25.2% identified earlier by the original cCDP. This combination of earlier detection and higher specificity yielded superior statistical separation (HR = 2.50, p < 0.0001 vs HR = 2.32, p < 0.0001) between progressor and improver arms at early trial timepoints (**Figure 4**).

**Table 3.** Digitally Enhanced cCDP events and original cCDP events in the progressor and improver arm.

|  | Progressor arm (n = 234) |  | Improver arm (n = 78) |  |
| --- | --- | --- | --- | --- |
|  | n | % | n | % |
| **Digitally Enhanced cCDP** |  |  |  |  |
| Censored | 91 | 38.9 | 67 | 85.9 |
| PwPMS with any progression event | 143 | 61.1 | 11 | 14.1 |
| EDSS events | 40 | 27.8<sup>a</sup> | 0 | 0<sup>b</sup> |
| 9HPT events | 25 | 17.4<sup>a</sup> | 6 | 54.6<sup>b</sup> |
| Digital gait events | 78 | 54.6<sup>a</sup> | 5 | 45.5<sup>b</sup> |
| **Original cCDP** |  |  |  |  |
| Censored | 88 | 37.6 | 65 | 83.3 |
| PwPMS with any progression event | 146 | 62.4 | 13 | 16.7 |
| EDSS events | 31 | 21.3<sup>c</sup> | 0 | 0<sup>d</sup> |
| 9HPT events | 35 | 24.0<sup>c</sup> | 6 | 46.2<sup>d</sup> |
| T25FW events | 80 | 54.8<sup>c</sup> | 7 | 53.9<sup>d</sup> |
| **Digitally Enhanced cCDP vs original cCDP** |  |  |  |  |
| New events detected | 28 | 12.0 | 4 | 5.1 |
| Events lost | 31 | 13.3 | 6 | 7.7 |
| **Progression events detected by both Digitally Enhanced and original cCDP** |  |  |  |  |
| PwPMS with progression events detected by both cCDP endpoints | 115 | 49.1 | 7 | 9.0 |
| Events detected earlier by the Digitally Enhanced cCDP | 49 | 42.6<sup>e</sup> | 2 | 28.65<sup>f</sup> |
| Events detected by both cCDP endpoints at the same time | 37 | 32.2<sup>e</sup> | 3 | 42.9<sup>f</sup> |
| Events detected later by the Digitally Enhanced cCDP | 29 | 25.2<sup>e</sup> | 2 | 28.6<sup>f</sup> |

<sup>a</sup>Percentage of PwPMS in the progressor arm with progression events detected by the Digitally Enhanced cCDP.
<sup>b</sup>Percentage of PwPMS in the improver arm with progression events detected by the Digitally Enhanced cCDP.

**Figure 4.**
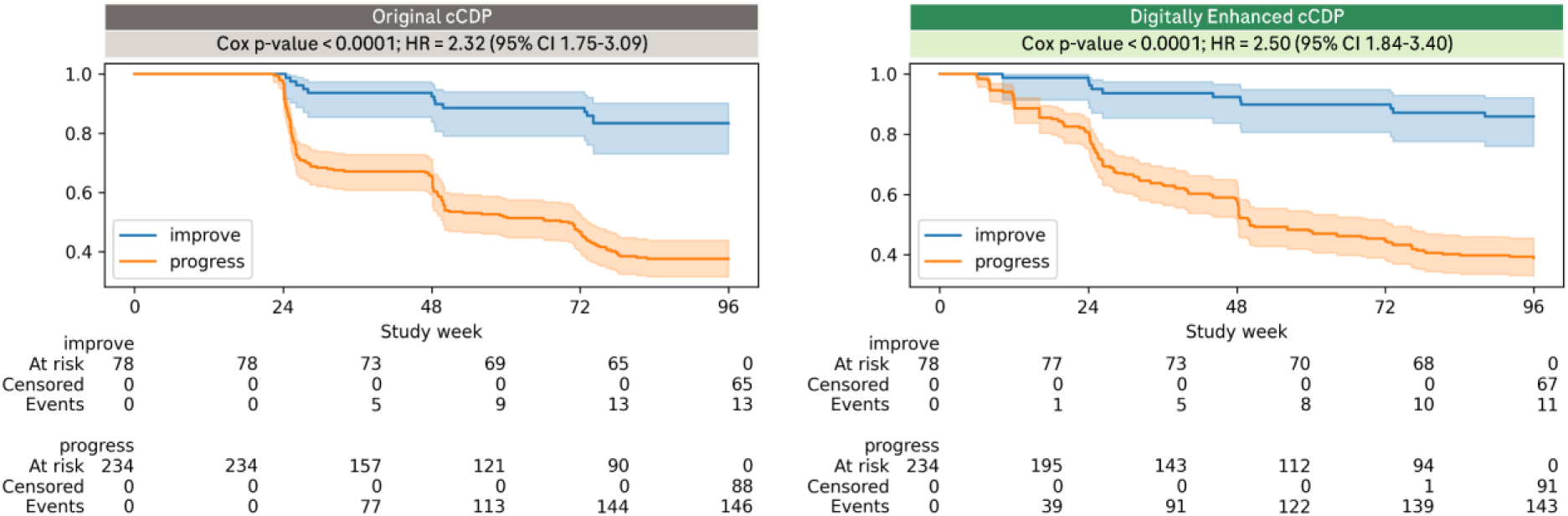
Survival curves of the Digitally Enhanced cCDP (A) and original cCDP (B). The sizes of the improver and progressor arms may vary due to differences in test adherence and data availability at the different time points. cCDP, composite Cumulative Disability Progression.

### Clinical Trial Efficiency and Sample Size Reductions

To evaluate potential operational efficiencies, sample sizes required to achieve 80% statistical power (*_α_* = 0.05) were compared across simulated trial durations (**Figure 5A**). The Digitally Enhanced cCDP achieved median sample size reductions of 24.9% (95% CI: 22.7– 27.1%) at Week 48, 21.2% (95% CI: 19.3–23.1%) at Week 72, and 14.6% (95% CI: 12.7–16.4%) at Week 96 relative to the original cCDP. Sensitivity analyses across various simulated effect sizes (*_k_* = 0 to *_k_* = 30 arm swaps) demonstrated that the Digitally Enhanced cCDP at Week 72 consistently achieved statistical power equivalent to or greater than the original cCDP at Week 96, demonstrating that trial observation periods can be shortened by 24 weeks (25%) without compromising statistical rigor (**Figure 5B**). Sample size reduction and ability to shorten observation periods by 24 weeks were confirmed for the sensitivity analysis employing a 24-week confirmation for digital events: 17.6% (95% CI: 15.1–20.1%) at Week 48, 14.8% (95% CI: 12.9–16.8%) at Week 72, and 4.1% (95% CI: 1.9–6.2%) at Week 96.

**Figure 5.**
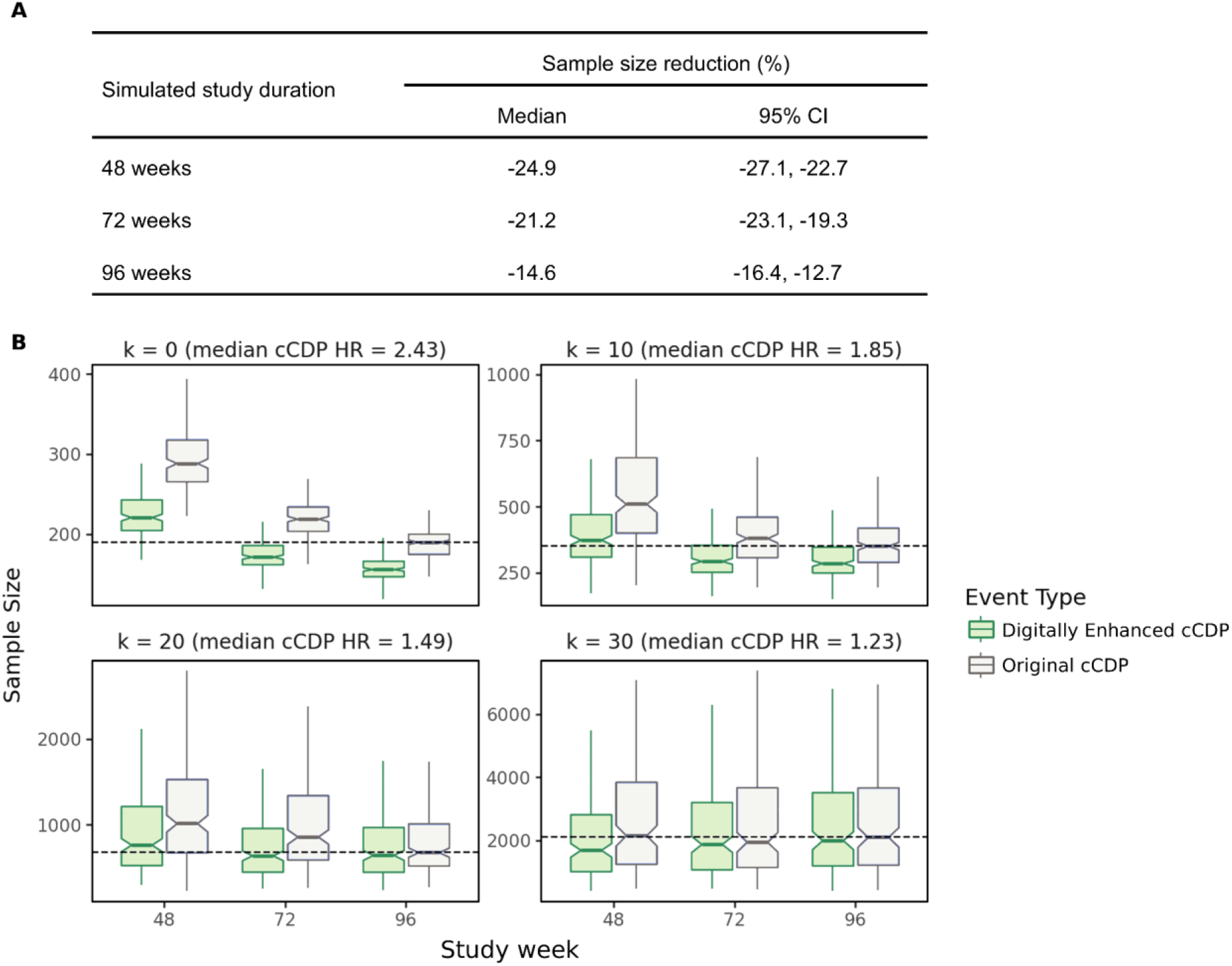
Sample size reduction achieved with the Digitally Enhanced cCDP. (A) When averaged across different simulated effect sizes, the Digitally Enhanced cCDP achieved a 14.6%–24.9% reduction in required sample size (with 80% statistical power) compared with the original cCDP. (B) Each panel shows a box plot of the sample size estimates (obtained from 200 sample replicates) needed to separate the improver and progressor arms at each time point for different values of *k* (i.e., different simulated effect sizes). To obtain equal sized arm, the larger arm was down-sampled, without replacement, to match the sample size of the smaller arm. The HR values are the median HR for cCDP over all replicates and time points for the indicated value of *k*. The notches on each box indicate the 95% confidence interval of the median. The dashed line indicates the median sample size required with the original cCDP at Week 96. Outliers are cropped to facilitate the graphical presentation. cCDP, composite Cumulative Disability Progression; HR, hazard ratio.

## Discussion

This study demonstrates that integrating remote, smartphone-based digital gait events into composite disability endpoints significantly improves clinical trial efficiency in progressive multiple sclerosis. By replacing the conventional in-clinic Timed 25-Foot Walk (T25FW) with continuous digital gait measures derived from a home-administered 2-minute walk test (2MWT), the Digitally Enhanced cCDP reduced required sample sizes by up to 25% at Week 48. Crucially, the digital endpoint achieved statistical power at Week 72 comparable to the traditional cCDP at Week 96, indicating that trial durations could be shortened by six months. These findings highlight a practical strategy for lowering operational costs, reducing participant burden, and more importantly accelerating therapeutic development in MS.

The superior performance of the Digitally Enhanced cCDP stems from the enhanced signal-to-noise ratio provided by frequent, objective remote assessments. ^5^ ^16^ In-clinic evaluations like the T25FW are episodic and highly susceptible to day-to-day performance variability, fatigue, and temporary symptoms, ^17^ ^18^ which can lead to misclassified progression events or regression to the mean. In contrast, aggregating daily home assessments over structured time windows averages out short-term fluctuations. To further prevent spurious events, the framework enforces a dual-threshold requirement where observed worsening must exceed both an adherence-adjusted Minimal Detectable Change (MDC) and a Minimal Clinically Important Difference (MCID). Here, the MDC acts as a statistical filter that dynamically lowers as patient adherence increases to confirm a change is not random noise, while the MCID ensures that change is large enough to impact real-world mobility. ^19^ ^20^ Requiring worsening to cross both thresholds increases specificity, effectively eliminating false-positive events in stable participants while enabling true progression to be more rapidly detected.

In addition to statistical benefits, digital gait monitoring offers greater ecological validity and patient relevance. ^21^ ^22^ Whereas the T25FW evaluates maximum walking speed over a brief distance in an artificial clinic setting, remote 2MWT measures capture real-world walking performance, including pace, endurance rhythm, and rest patterns. ^13^ The inclusion of Loss of Perceived Walking Safety (LPWS) addresses a critical gap in traditional trial design. In standard clinical trials, participants unable to perform in-clinic gait tests due to severe disability are typically assigned arbitrary ceiling scores (e.g., 180 seconds on T25FW ^23^). By tracking skipped tests driven by self-reported safety concerns, LPWS captures functional deterioration and in individuals with advanced disability who might otherwise be unmeasurable on speed measures alone.

Several study limitations should be noted. First, performance was evaluated using simulated two-arm trials derived from long-term EDSS trajectories within a single-arm study (CONSONANCE). While sensitivity analyses confirmed that sample size reductions persist across a broad range of effect sizes, prospective validation in randomized, controlled trials is required. Second, protocol adherence was unconstrained in this real-world setting. This factor, alongside administrative censoring, explains the slight reduction in sample size benefit during the 24-week confirmation sensitivity analyses, as longer confirmation periods make successful event confirmation less likely (especially at later time points). However, because higher adherence decreases MDC noise thresholds, enforcing stricter adherence in future protocols would only serve to further enhance sensitivity and event detection power.

In conclusion, our framework successfully translates high-frequency continuous and categorical digital measures into validated time-to-event endpoints. By replacing episodic clinic assessments with robust digital gait measures, the Digitally Enhanced cCDP offers a validated pathway to design smaller, shorter, and more ecologically valid clinical trials in multiple sclerosis. Beyond ambulatory monitoring, this modular framework provides a template for integrating digital assessments across other functional domains, including upper limb function and cognition, to refine endpoint sensitivity in future neurological disease trials.

## Supporting information

Institutional Review Boards and Ethics Committees for the Consonance Trial (NCT03523858)

## Declarations

## Acknowledgments

The authors would like to thank all study participants and their families. They also thank Mattia Zanon and Guy Bogaarts, both of F. Hoffmann-La Roche Ltd, for the contributions to the design of the framework for detecting progression events using digital measures.

Editorial and writing support was provided by Sven Holm, PhD, contractor for F. Hoffmann-La Roche Ltd. The authors confirm that all intellectual content, data interpretation, and conclusions were generated by the authors. This research was funded by F. Hoffmann-La Roche Ltd, Basel, Switzerland.

## CRediT Authorship Contribution Statement

**AF** Data Curation, Software, Formal Analysis, Methodology, Visualization, Writing – Original Draft Preparation, Writing – Review & Editing

**DS** Data Curation, Software, Formal Analysis, Methodology, Visualization, Writing – Original Draft Preparation, Writing – Review & Editing

**CS** Data Curation, Software, Formal Analysis, Methodology, Visualization, Writing – Review & Editing

**JR** Formal Analysis, Methodology, Visualization, Writing – Review & Editing

**LL** Conceptualization, Investigation, Writing – Review & Editing

**AK** Supervision, Project Administration, Writing – Review & Editing

**JO** Conceptualization, Writing – Review & Editing

**JL** Conceptualization, Writing – Review & Editing

**GC** Conceptualization, Writing – Review & Editing

**HB** Conceptualization, Investigation, Writing – Review & Editing

**LC** Conceptualization, Methodology, Funding Acquisition, Supervision, Writing – Review & Editing

**MR** Conceptualization, Formal Analysis, Methodology, Project Administration, Supervision, Visualization, Writing – Original Draft Preparation, Writing – Review & Editing

## Competing Interests

**AF** is a contractor for F. Hoffmann-La Roche Ltd.

**DS** is a consultant for F. Hoffmann-La Roche Ltd.

**CS** is an employee of F. Hoffmann-La Roche Ltd

**JR** is a consultant for F. Hoffmann-La Roche Ltd.

**LL** has received compensation for consulting services from Almirall, EXCEMED, F. Hoffmann-La Roche Ltd, Merck, Novartis, Biogen, Janssen-Cilag and Bristol Myers Squibb.

**AK** is an employee of and shareholder in F. Hoffmann-La Roche Ltd.

**JO** is an employee of and shareholder in F. Hoffmann-La Roche Ltd.

**JL**’s former institution (University Hospital Basel) has received grants from Innosuisse - Swiss Innovation Agency, the MSBase Foundation, Novartis, and Biogen and honoraria for advisory boards and/or speaking fees from Novartis, Roche and Teva exclusively used for funding research. He received travel compensation from Novartis and Bristol Myers Squibb.

**GC** has received compensation for participation in data and safety monitoring boards for Applied Therapeutics, AI Therapeutics, AMO Pharma, Argenx, _AstraZeneca_, Avexis Pharmaceuticals, Bristol Meyers Squibb, CSL Behring, Cynata Therapeutics, _DiaMedica_ Therapeutics, Horizon Pharmaceuticals, Immunic, Inhibrix, Karuna Therapeutics, Kezar Life Sciences, Medtronic, Merck, Meiji Seika Pharma, Mitsubishi Tanabe Pharma Holdings, Prothena Biosciences, Novartis, Pipeline Therapeutics (Contineum), Regeneron, Sanofi-Aventis, Teva Pharmaceuticals, United _BioSource_, and University of Texas Southwestern, and in consulting for Alexion, Antisense Therapeutics–Percheron, Avotres, Biogen, Clene Nanomedicine, Clinical Trial Solutions, Endra Life Sciences, Cognito Therapeutics, F Hoffman-La Roche, Genentech, Genzyme, Hoya Corporation, Immunic, Immunosis Pty, Klein-Buendel Incorporated, Kyverna Therapeutics, Linical, Merck–Serono, Noema, Neurogenesis, Perception Neurosciences, Protalix Biotherapeutics, Regeneron, Revelstone Consulting, SAB Biotherapeutics, Sapience Therapeutics, Scott & Scott, and Tenmile; has received institutional grants from University of Alabama at Birmingham, AL, USA; and holds non-financial leadership or fiduciary roles in the Birmingham Jewish Federation, Birmingham Jewish Foundation, and Graffman Endowment Committee.

**HB** is an employee of Monash University and board member and managing director of MSBase Foundation. He has received conference travel compensation from Novartis and Merck. His institution received honoraria for talks and lectures from Merck, F. Hoffmann-La Roche Ltd, Novartis. He has received personal compensation for talks from Neuraxpharm. His institution has received compensation for activities in study steering committees, advisory boards, and discussion forums from F. Hoffmann-La Roche Ltd, Novartis, and Merck and research grants from F. Hoffmann-La Roche Ltd., Argenx, Novartis, Merck, UCB Pharma, the Medical Research Future Fund Australia, NHMRC Australia, Trish Foundation, MS Australia, and Monash University.

**LC** is an employee of and shareholder in F. Hoffmann-La Roche Ltd.

**MR** is a contractor for F. Hoffmann-La Roche Ltd.

## Data Availability

Up-to-date details on Roche’s Global Policy on Sharing of Clinical Study Information and how to request access to related clinical study documents are available on https://go.roche.com/data_sharing. Request for the data underlying this publication requires a detailed, hypothesis-driven, statistical analysis plan that is collaboratively developed by the requestor and company subject matter experts. Such requests should be directed to for consideration. Anonymized records for individual patients across more than one data source external to Roche cannot, and should not, be linked due to a potential increase in risk of patient reidentification.

## Supplementary tables and figures

**Supplementary Table S1.** Summary of psychometric properties of digital measures used in the event-based endpoint.

|  | Step Velocity Scaled to Walking Time | Step Duration |
| --- | --- | --- |
| Concurrent validity against reference systems (Gait Up IMU sensors): ICC(3,1) for absolute agreement, median (IQR) | 0.93 (0.90 – 0.95)<sup>a</sup> | 0.93 (0.89 – 0.96) |
| Test-retest reliability: ICC(2,1) for absolute agreement, median (IQR) |  |  |
| Single test | 0.87 (0.84 – 0.90) | 0.87 (0.81 – 0.90) |
| 3 aggregated tests | 0.95 (0.94 – 0.96) | 0.95 (0.93 – 0.96) |
| 6 aggregated tests | 0.98 (0.97 – 0.98) | 0.98 (0.97 – 0.98) |
| Convergent validity: Spearman rank-order correlation, $\rho$ (IQR) | | |
| T25FW | 0.80 (0.72 – 0.87) | -0.71 (-0.80 to -0.59) |
| EDSS | -0.69 (-0.78 to -0.56) | 0.64 (0.49 – 0.75) |
| Ambulation Score | -0.67 (-0.77 to -0.53) | 0.66 (0.52 – 0.77) |
| MSWS-12 | -0.70 (-0.80 to -0.58) | 0.64 (0.5 – 0.75) |
| Known-groups validity |  |  |
| EDSS 0 – 3.5, median (IQR) | 1.31 (1.17 – 1.40) | 0.52 (0.48 – 0.57) |
| EDSS 4.0 – 5.5, median (IQR) | 1.12 (0.80 – 1.26) | 0.55 (0.52 – 0.60) |
| EDSS 6.0 – 6.5, median (IQR) | 0.62 (0.45 – 0.84) | 0.70 (0.61 – 0.81) |
| p-value EDSS 0 – 3.5 vs EDSS 4.0 – 5.5 | $p < 0.01$ | $p = 0.08$ |
| p-value EDSS 4.0 – 5.5 vs EDSS 6.0 – 6.5 | $p < 0.01$ | $p < 0.01$ |
| MDC for n number of tests within an aggregation window |  |  |
| MDC for < 7 tests within an aggregation window | $MDC_{97.5}(n) = 0.4288 * n^{-0.4936}$ | $MDC_{99}(n) = 0.1964 * n^{-0.5989}$ |
| MDC for $\geq 7$ tests within an aggregation window | $MDC_{97.5} = 0.1641$ | $MDC_{99} = 0.0613$ |
| MCID for progression | -0.18 | +0.06 |

<sup>a</sup>ICC(3,1) of unscaled Step Velocity was used instead, because the reference measurement system does not provide a version scaled to walking time. EDSS; Expanded Disability Status Scale; ICC, intraclass correlation coefficient; IQR, interquartile range; MCID, minimal clinically important difference; MDC, minimal detectable change; MSWS-12, 12-item Multiple Sclerosis Walking Scale; T25FW, Timed 25-Foot Walk.

**Supplementary Table S2.** Baseline demographics and disease characteristics: GaitLab study.

|  | **GaitLab (n = 96 PwMS)** |
| --- | --- |
| Age, years, median (IQR) | 56.0 (46.8 – 63.2) |
| Female, n (%) | 59 (61.5) |
| Height, m, median (IQR) | 1.7 (1.6 – 1.8) |
| Weight, kg, median (IQR) | 75.0 (64.8 – 93.0) |
| MS type, n (%) |  |
| Relapsing-remitting | 67 (69.8) |
| Secondary progressive | 16 (16.7) |
| Primary progressive | 13 (13.5) |
| Time since MS symptom onset, years, median (IQR) | 18 (11 – 27) |
| T25FW, s, median (IQR) | 5.4 (4.5 – 8.4) |
| Ambulation Score, median (IQR) | 3.5 (1.0 – 5.0) |
| MSWS-12 v2 transformed total score, median (IQR) | 47.0 (21.0 – 73.0) |
| EDSS |  |
| Median (IQR) | 5.3 (4.0 – 6.0) |
| 0 to 3.5, n (%) | 23 (24.0) |
| 4.0 to 5.5, n (%) | 35 (36.5) |
| 6.0 to 6.5, n (%) | 38 (39.6) |

EDSS; Expanded Disability Status Scale; IQR, interquartile range; MS, multiple sclerosis; MSWS-12, 12-item Multiple Sclerosis Walking Scale; N/A, not applicable; PwMS, people with multiple sclerosis; T25FW, Timed 25-Foot Walk.

**Supplementary Figure S1.**
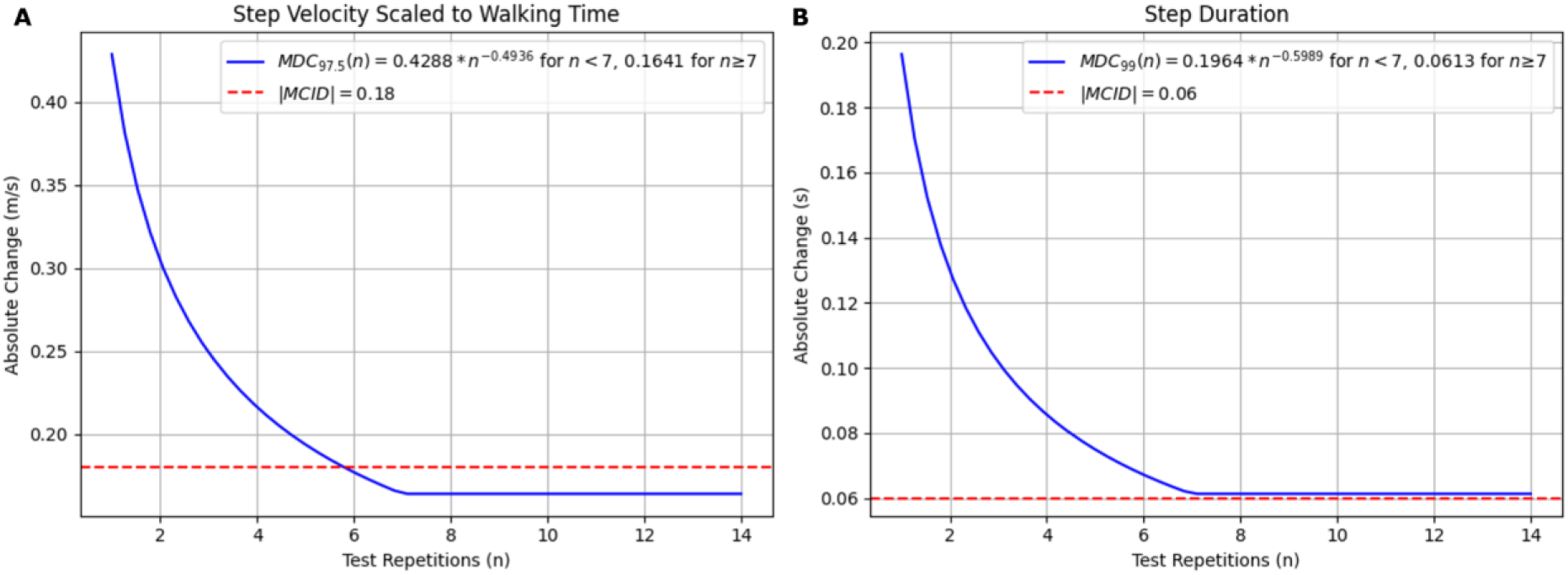
The relationship between MCID and MDC is shown as a function of test executions within a window for both Step Velocity Scaled to Walking Time (A) and Step Duration (B). MDC predominantly influences decisions when the number of test repetitions is low, whereas MCID becomes the dominant factor once test repetitions exceed 7. In the GaitLab study, MDC could only be estimated up to (n = 7), so the curve was conservatively truncated at this point.

