## Supplementary material for "Remote ambulation monitoring enhances composite disability measurement in MS to deliver reduced trial sample size": Institutional Review Boards and Ethics Committees for the Consonance Trial (NCT03523858)

| Country | Ethics Committee / Institutional Review Board (IRB) Name |
|---|---|
| Bosnia and Herzegovina | Ethics Committee EC University Clinical Center Tuzla |
| Brazil | CEP do Instituto de Neurologia de Curitiba S/C Ltda. |
| | Comissão de Ética para Análise de Projetos de Pesquisa - CAPPesq |
| | Comitê de Ética em Pesquisa com Seres Humanos; Hospital das Clínicas de Ribeirão Preto |
| | Comitê de Ética em Pesquisa da PUCRS |
| Canada | Alberta Health Serivces REB |
| | Comitéd'éthique de la recherche, CHUM |
| | Fraser Health Research Ethics Board |
| | IRB Services |
| | St Mike's Hospital Research Ethics Board |
| | University of Saskatchewan Biomedical Research Ethics Board |
| | Western University Health Science Research Ethics Board |
| Colombia | Comite De Etica En Investigacion Fundacion Universitaria Sanitas |
| | Comité de Investigación y Etica |
| | Comite De Investigaciones Y Etica |
| Costa Rica | Comite Etico Cientifico Universidad de Ciencias Medicas |
| Czech Republic | Eticka Komise Fakultni nemocnice Olomouc a Lekarske fakulty UP v Olomouci |
| | Eticka komise Vseobecne fakultni nemocnice v Praze |
| Denmark | De Videnskabsetiske Komitéer for Region Hovedstade |
| Egypt | Alexandria University Research Thics committee |
| | Faculty of Medicine-Ain Shams University |
| France | CPP Ile de France VI-Pitié Salpêtrière |
| Germany | Ethikkommission an der Medizinischen Fakultät Ernst-Moritz-Arndt-Universität Greifswald |
| Guatemala | Comité de Ética Independiente Zugueme |
| Hungary | Egeszsegugyi Tudomanyos Tanacs - Klinikai Farmakologiai Etikai Bizottsaga |
| | Jahn Ferenc Dél-pesti Kórház és Rendelőintézet; Intézményi Kutatásetikai Bizottság |
| | Semmelweis Univ. Reg. and Instit.Committe od S&RE |
| | University of Szeged Committee |
| | Uzsoki Utcai Korhaz Etikai Bizottsaga |
| Ireland | National Research Ethics Committee (NREC) |
| Italy | Comitato Etico Territoriale Lombardia 1; presso IRCCS Ospedale San Raffaele |
| Lebanon | American University of Beirut - Medical Center; IRB |
| Mexico | Comite de Etica en Investigacion del Hospital General de Mexico, Doctor Eduardo Liceaga |
| | Comite de Etica en Investigacion del Medica Sur, S.A.B. de C.V |
| | Comité de Ética en Investigación Sanatorio Alcocer Pozo S.A de C.V. |
| Morocco | Comité d'Ethique de la Recherche Biomédicale; Faculté de Médecine et de Pharmacie de Rabat |
| Netherlands | Stichting BEBO |
| | Stichting Beoordeling Ethiek Biomedisch Onderzoek (BEBO) |

| Country | Ethics Committee / Institutional Review Board (IRB) Name |
|---|---|
| | Stichting Beoordeling Ethiek Biomedisch Onderzoek (Stichting BEBO); Ambtelijk Secretariaat |
| Panama | Comite de Bioetica de Investigacion del Hospital Punta Pacifica |
| Poland | Akademia Medyczna im. K. Marcinkowskiego |
| | Komisja Bioetyczna przy Okręgowej Izbie Lekarskiej w Łodzi |
| | Komisja Bioetyczna Slaskiego Uniwersytetu Medycznego |
| | Komisja Bioetyczna Uniwersytetu Jagiellońskiego |
| | Komisja Bioetyczna Uniwersytetu Medycznego w Białymstoku |
| | Komisja Bioetyczna WIM Wojskowy Instytut Medyczny |
| | Okręgowa Izba Lekarska |
| Russian Federation | EC of Jusupovskaya Hospital |
| | EC of National Center of Social Significant Disease |
| | EC of Scientific Neurology Center; Neurological department #6 |
| | Moscow State Independent Ethics Committe |
| | NEC of VLADIMIRSKIY REGIONAL SCIENTIFIC RESEARCH INST. |
| Spain | CEIC Hospital Vall D'Hebron |
| United Arab Emirates | Cleveland Clinic Abu Dhabi |
| | Rashid hospital |
| United States | Committee on Human Research (CHR) |
| | Johns Hopkins Medicine Institutional Review Board |
| | University of Pennsylvania; Office of Regulatory Affairs-IRB |
